# Balloon guide catheter versus different sizes of non-balloon guide catheter; a MR CLEAN Registry analysis

**DOI:** 10.1101/2023.07.07.23292400

**Authors:** Robrecht R.M.M. Knapen, Robert-Jan B. Goldhoorn, Jeannette Hofmeijer, Geert J. Lycklamaà Nijeholt, René van den Berg, Ido R. van den Wijngaard, Robert J. van Oostenbrugge, Wim H. van Zwam, Christiaan van der Leij, MR CLEAN Registry Investigators

## Abstract

**Introduction:** Balloon guide catheters (BGC) are used to prevent distal emboli during endovascular treatment (EVT) for acute ischemic stroke. Although literature reports benefit of BGC, these are not universally used and randomized head-to-head comparisons are lacking. This study compared functional, safety, and technical outcomes between patients treated with different sizes non-BGC and with BGC during EVT in a nationwide prospective multicenter registry.

**Methods:** Patients from the MR CLEAN Registry (2014–2018), who underwent EVT with a 5-7French (Fr) non-BGC, a 8-9Fr non-BGC, or a 8-9Fr BGC were included. Primary outcome was the modified Rankin Scale (mRS) score at 90 days, secondary outcomes included procedure time and first-attempt successful reperfusion (eTICI ≥ 2C). Treatment-effect modification and subgroups were analyzed according to first-line thrombectomy technique: stent retriever (SR) or direct aspiration (ASP).

**Results:** In total 2808 patients were included, and 1671 (60%) were treated with 8-9Fr BGC. Overall, no significant differences in clinical outcome were seen between non-BGC and BGC groups. The 8-9Fr non-BGC was associated with lower first-attempt successful revascularization rates compared to BGC (aOR:0.76, 95%CI:0.59-0.998), the 5-7 Fr non-BCG was not. Regression analysis showed a significant interaction between BGC use and device type. In the subgroup with SR as first-line technique, 90 day mRS scores were significantly higher in the 8-9Fr non-BGC group compared with BCG (acOR:0.77; 95%CI:0.59-0.996), but not in the 5-7Fr non-BCG. Direct aspiration combined with 5-7Fr non-BGC resulted in higher first-attempt rates compared to BGC (aOR:1.75; 95%CI:1.16-2.63).

**Conclusion:** This large prospective multicenter registry showed no differences in clinical outcome between patients treated with 5-7Fr non-BGC, 8-9Fr non-BGC, and 8-9Fr BGC. Subgroup analyses, however, suggest that BCG outperforms the non-BGC when SR is used as first-line technique.

## Introduction

The use of a balloon guide catheter (BGC) during endovascular treatment (EVT) of acute ischemic stroke (AIS) is a well-known technique for achieving flow arrest, in order to avoid potential clot fragmentation with distal emboli.(1) Use of BGC is associated with shorter procedure time, higher successful reperfusion rates, and better functional outcomes when compared to a non-BGC approach, most of these studies are based on anterior circulation occlusions.(2–5).

Two studies compared, in addition, the use of a non-BGC and BCG in combination with stent retriever thrombectomy. The Solitaire or Trevo stent retriever showed higher functional outcomes and higher reperfusion rates when a BGC was used.(6, 7) Still, many procedures are performed without a BGC. A survey questionnaire showed that only 25% of treating physicians routinely used a BGC.(8) Arguments against BGC are higher costs, its rigidity, and the need for larger sheaths. On the other side, BGCs are rapidly evolving and the additional costs are low compared to the overall costs.(9).

The primary aim of this study is to investigate clinical, technical, and safety outcomes between a 5-7Fr non-BGC, a 8-9Fr non-BGC, and a 8-9Fr BGC approach in a nationwide registry of acute ischemic stroke patients treated with EVT. Secondary aim is to compare clinical and technical outcome between the three guide catheters when combined with either stent retriever thrombectomy or direct aspiration thrombectomy as first-line technique.

## Methods

### Design

For this study, we used data from the Multicenter Randomized Clinical Trial of Endovascular Treatment for Acute Ischemic Stroke in The Netherlands (MR CLEAN) Registry. All AIS patients who underwent EVT because of an intracranial large vessel occlusion between March 2014 and December 2018, were included in this registry.

The MR CLEAN Registry study protocol was granted with the permission to carry out the study as a registry after evaluating by the medical ethics committee of the Erasmus University Medical Center (MEC-2014-235). The committee waived the need for obtaining informed consent.

This study was conducted using the STROBE guidelines. The corresponding author takes responsibility for its integrity and data analysis and had full access to all study data. Due to legislative issues on patient privacy source data will not be made available. On reasonable request to the corresponding author, detailed statistical analyses will be made available.

### Participants

For this study, we included patients older than 18 years treated with EVT within 6.5 hours after the start of stroke symptoms due to intracranial large vessel occlusion in the anterior circulation (intracranial carotid artery and middle cerebral artery). When insufficient data were available regarding the used (balloon) guide catheter patients were excluded.

### Treatment

A BGC was registered when a 8-9Fr guide catheter and a balloon were registered on the intervention form. The choice of a BGC was made by the local treating physician. No data were available regarding whether the balloon was inflated, and if flow reversal was achieved with the guide catheter.

### Outcomes

The primary outcome was the modified Rankin Scale (mRS) score at 90 days of follow-up, ranging from 0 (no symptoms) to 6 (death). Secondary functional outcome measurements were excellent (defined as mRS score 0-1) and favorable (defined as mRS score 0-2) functional outcome, and National Institutes of Health Stroke Scale (NIHSS) score 24-48 hours after EVT. Technical outcomes included procedure duration, reperfusion grade, and first-attempt successful reperfusion, whereas safety outcomes included ischemic stroke progression and occurrence of symptomatic intracranial hemorrhage (sICH).

Reperfusion grade was based on the extended Thrombolysis In Cerebral Infarction (eTICI), a scale ranging from 0 (no reperfusion) to 3 (complete reperfusion). Successful reperfusion was defined by eTICI ≥ 2B, excellent reperfusion by eTICI ≥ 2C, and complete reperfusion by eTICI 3. First-attempt successful reperfusion was defined as eTICI ≥ 2C after one attempt. Thrombus in another territory was scored as a remaining occlusion, which did not match the target occlusion and had changed to another territory or changed to a more proximal location. An independent core lab consisted of two neuroradiologists and six interventional (neuro)radiologists assessed all the imaging separately, while they were blinded for all clinical findings or findings on other imaging modalities. Stroke progression was defined when a patient scored at least 4 points higher on the NIHSS. sICH was defined as an intracranial hemorrhage related to the clinical deterioration according to the Heidelberg criteria in combination with a neurologic deterioration of an increase of ≥ 4 points on the NIHSS score. An adverse events committee scored the sICHs.

### Statistical analysis

Baseline characteristics were presented with standard statistics. To compare patients treated with different guide catheters, we used a χ^2^ test for binary or ordinal outcomes, whereas an independent-samples t-test was used for continuous parameters, after checking for normality of distribution using plots and Shapiro-Wilk test.

For the primary outcome, multiple ordinal regression analysis was used to compare the effect of the different guide catheters on the mRS at 90 days follow-up with BGC as comparator. Secondary outcomes were analyzed with multiple ordinal, binary, or linear regression analyses as appropriate. Odds ratios (OR) or beta estimates with 95% confidence intervals were used to present the regression model results. Variables for adjusting the regression models were chosen based on literature and baseline characteristics differences. These variables were age, sex, baseline NIHSS, pre-stroke mRS, time between start symptoms and start EVT, baseline collateral score, atrial fibrillation, and location of the occlusion.

R Studio (version 1.3.1093) was used to perform all statistics. The level of significance was set at 0.05.

### Subgroup analyses

We studied two pre-defined subgroups: depending on location of the occlusion and which first-line technique was used. The interaction with effect was estimated for these subgroups. Hereto, two interaction terms were added, one between the location of the occlusion and the guide catheter, and one between the first-line technique (stent retriever thrombectomy or direct aspiration thrombectomy) and the guide catheter. Patients treated with stent retriever combined with direct aspiration thrombectomy as first-line technique, were classified as treated with stent retriever thrombectomy. When treatment interaction was significant, subgroup analyses were performed to evaluate the effect on the mRS score, otherwise exploratory subgroup analyses were given. Regardless of the sample size, the same adjustments were made as for the regression analyses.

### Missing data

All descriptive analyses were performed with original data. For the regression analyses, missing data were replaced with data obtained from multiple imputations. Multiple imputations were performed with predefined variables as predictors, for the complete list see supplemental S1.

## Results

A total of 2808 patients (cohort March 2014 till December 2018,) were included in this study, of which 1671 (60%) were treated with a BGC (figure 1). At baseline, patients treated using a BGC more often had atrial fibrillation compared to the non-BGC groups. The time between the start of the symptoms or the moment of last seen well and groin puncture was the longest in the 5-7Fr non-BGC (mean 203 minutes). Distal intracranial internal carotid artery occlusions were most common in the 8-9Fr non-BGC group, while occlusions in segment M1 of the middle cerebral artery were more common in the 5-7Fr non-BGC group. All baseline characteristics are described in table 1.

**Figure 1.**
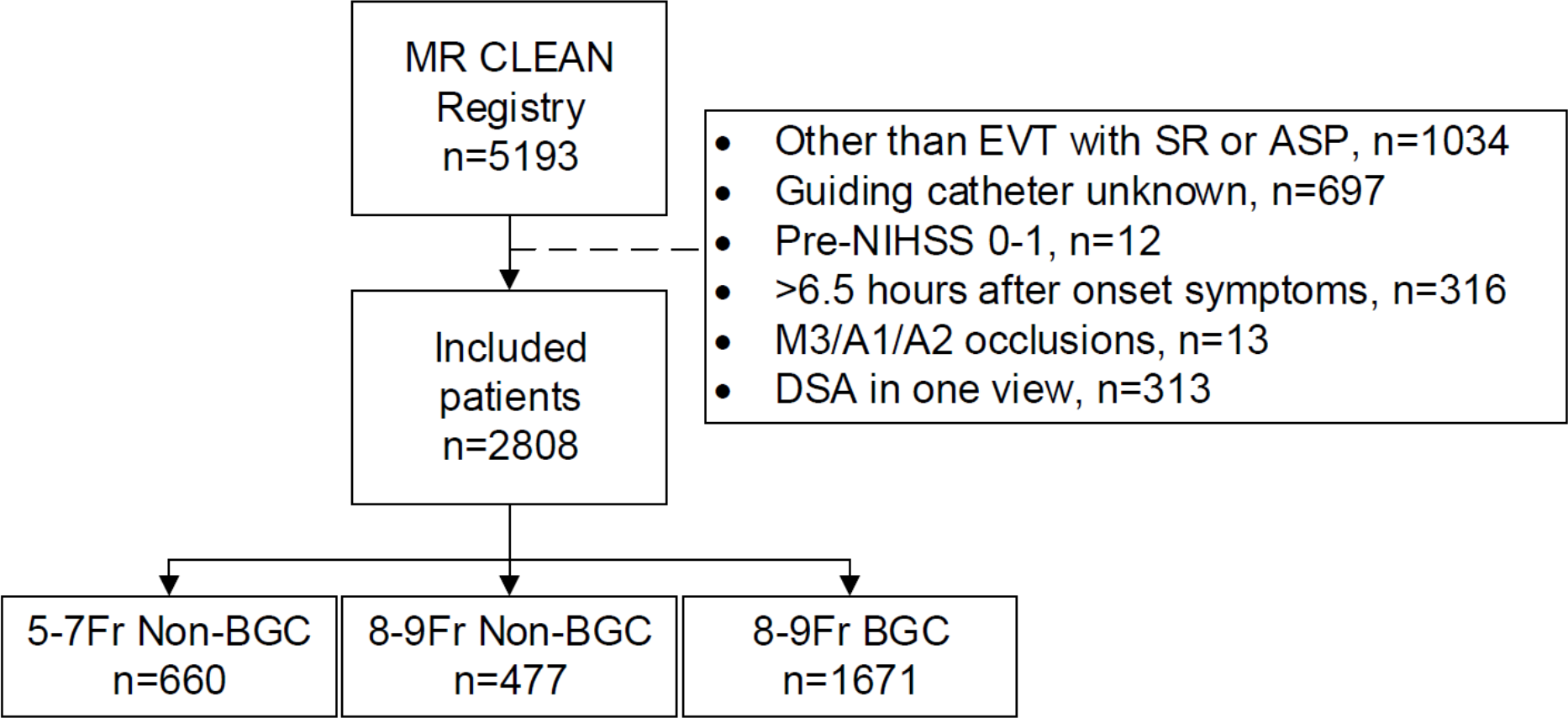
Flowchart of included patients. MR CLEAN, Multicenter Randomized Clinical Trial of Endovascular Treatment for Acute Ischemic Stroke in The Netherlands (MR CLEAN); EVT, endovascular treatment; SR, stent retriever; ASP, aspiration; NIHSS, National Institutes of Health Stroke Scale; DSA, digital subtraction angiography; Fr, French; BGC, balloon guide catheter.

**Table 1:**
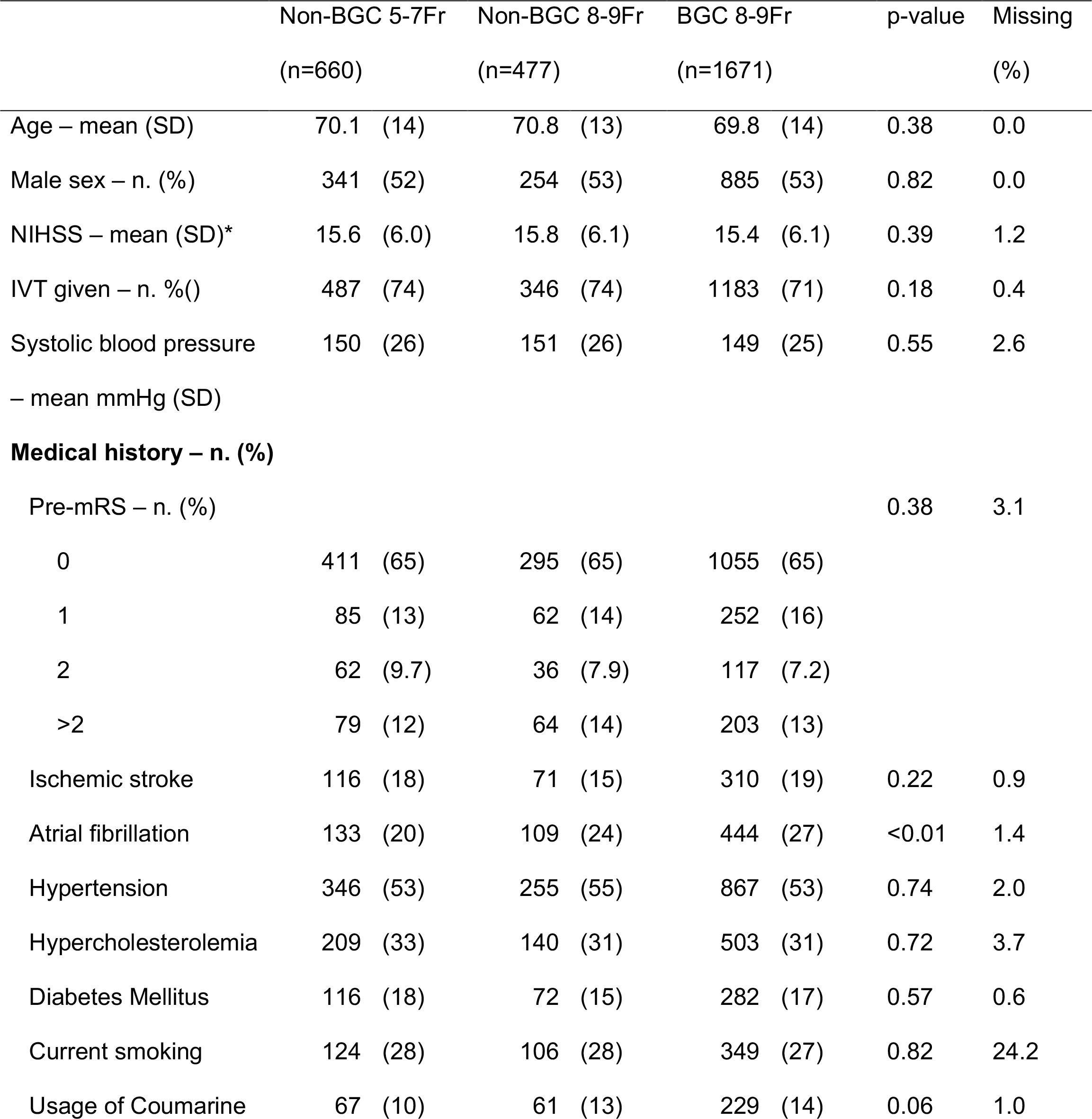
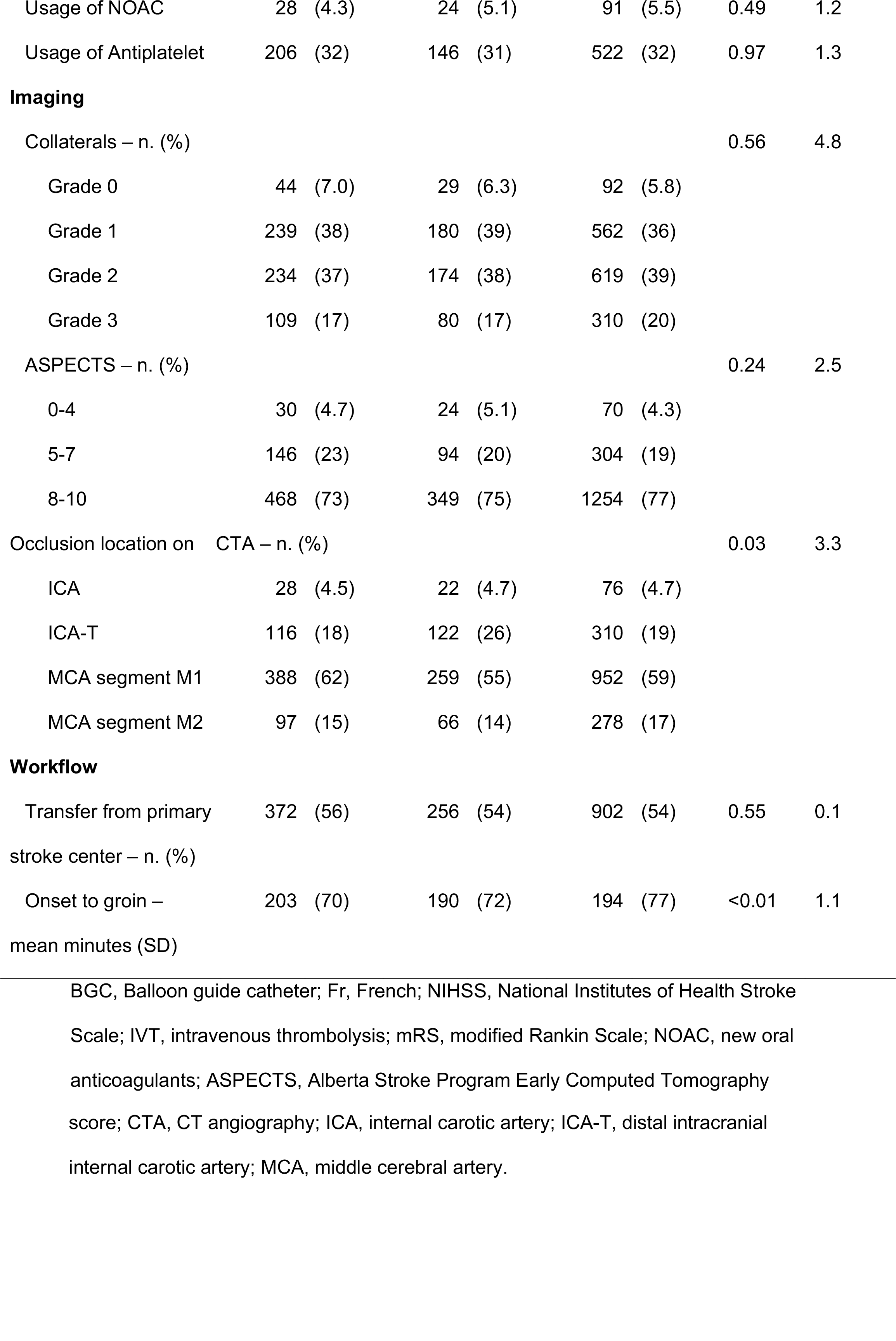
Baseline characteristics of the patients treated with a BGC compared to patients treated with a non-BGC.

### Functional outcome

There was no significant difference in outcome according to the 90 day mRS between the BGC and the 5-7Fr non-BGC group (adjusted common (ac)OR:0.94, 95%CI:0.78-1.12; figure 2), or between the BGC group and the 8-9Fr non-BGC group (acOR:0.96, 95%CI:0.79-1.18).

**Figure 2.**
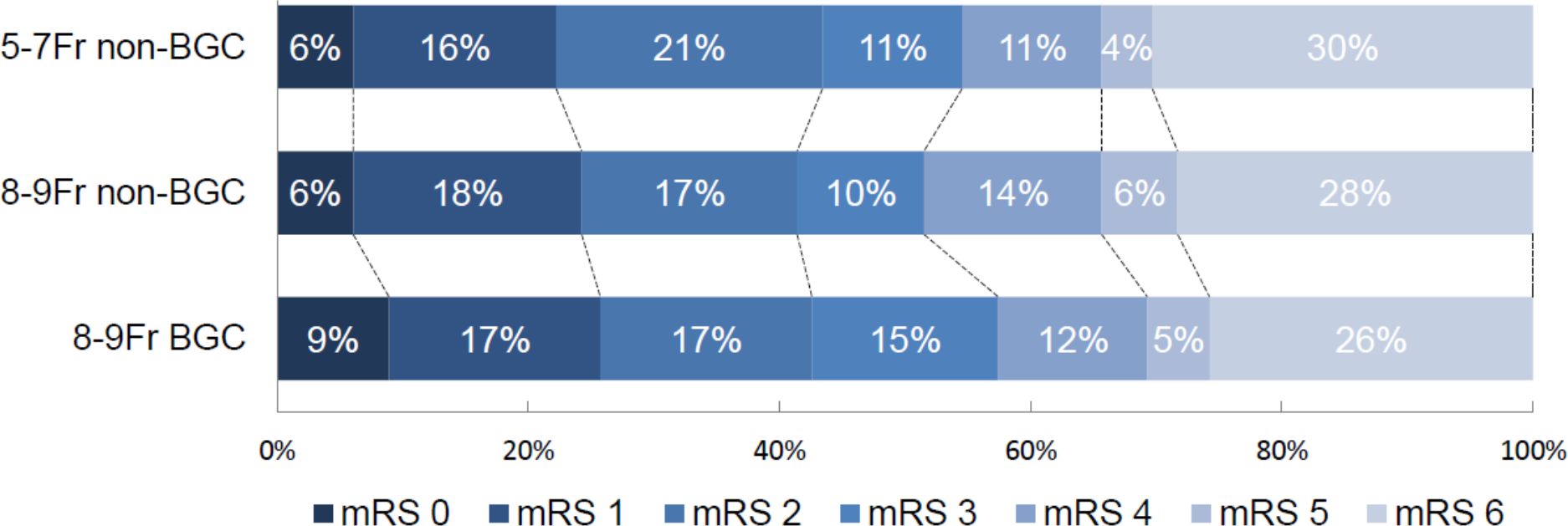
Distribution of the modified Rankin Scale between the use of a 5-7Fr non-BGC, 8-9Fr non-BGC and 8-9Fr BGC. Multiple logistic regression with BGC as comparator with adjustment showed no significant difference in mRS score after 90 days between the 5-7Fr non-BGC (acOR:0.94, 95%CI:0.78-1.12) and the 8-9Fr non-BGC (acOR:0.96, 95%CI:0.79-1.18).

Regression analyses showed no differences in favorable functional outcome between the BGC and the 5-7Fr- and 8-9Fr non-BGC (aOR:1.20, 95%CI:0.95–1.51, and aOR:1.08, 95%CI:0.83-1.40, respectively). Additionally, no differences were seen in NIHSS improvement with ≥ 4 points after 24-48 hours between 5-7Fr non-BGC versus BGC (aOR:1.04, 95%CI:0.84-1.28) and 8-9Fr non-BGC and the BGC (aOR:0.81, 95%CI:0.64-1.02).

### Technical outcome

The 5-7Fr non-BGC, 8-9Fr non-BGC and the 8-9Fr BGC did not differ in successful reperfusion rates (77%, 76% and 77% respectively) (Table 2). The use of a 5-7Fr non-BGC resulted in higher complete reperfusion rates (44% versus 40%, aOR:1.26, 95%CI:1.04–1.53; Table 3) and shorter procedure time (mean 54 minutes versus 57 minutes; aβ:-3.71, 95%CI:-6.79 to −0.63; Table 3) as compared with the BGC group. First-attempt successful revascularization rate was lower in the 8-9Fr non-BGC group compared to the BGC group (39% versus 46%, aOR:0.76, 95%CI:0.59-0.998; Table 3).

**Table 2.** Outcomes between patients treated with a BGC and patients treated with a non-BGC.

|  | Non-BGC 5-7Fr<br>(n=660) | Non-BGC 8-9Fr<br>(n=477) | BGC 8-9Fr<br>(n=1671) | p-value |
| --- | --- | --- | --- | --- |
| mRS at 90 days – n. (%) |  |  |  | 0.02 |
| 0 | 35/580 (6.0) | 28/436 (6.4) | 132/1544 (8.6) |  |
| 1 | 94/580 (16) | 78/436 (18) | 267/1544 (17) |  |
| 2 | 124/580 (21) | 75/436 (17) | 269/1544 (17) |  |
| 3 | 65/580 (11) | 44/436 (10) | 226/1544 (15) |  |
| 4 | 63/580 (11) | 61/436 (14) | 182/1544 (12) |  |
| 5 | 24/580 (4.1) | 27/436 (6.2) | 75/1544 (4.9) |  |
| 6 | 175/580 (30) | 123/436 (28) | 393/1544 (26) |  |
| mRS score 0-1 – n. (%) | 129/580 (22) | 106/436 (24) | 399/1544 (26) | 0.22 |
| mRS score 0-2 – n.(%) | 253/580 (44) | 181/436 (42) | 668/1544 (43) | 0.77 |
| Successful reperfusion | 489/634 (77) | 354/464 (76) | 1263/1633 (77) | 0.89 |
| (eTICI ≥2B) – n. (%) | | | | |
| Excellent reperfusion | 351/634 (55) | 243/464 (52) | 865/1633 (53) | 0.52 |
| (eTICI ≥2C) – n. (%) | | | | |
| Complete reperfusion | 280/634 (44) | 176/464 (38) | 647/1633 (40) | 0.07 |
| (eTICI = 3) – n. (%) |  |  |  |  |
| Symptomatic ICH – n. (%) | 36 (5.5) | 26 (5.5) | 102 (6.1) | 0.77 |
| Ischemic stroke | 54 (8.2) | 41 (8.6) | 136 (8.1) | 0.95 |
| progression – n. (%) |  |  |  |  |
| Mortality at 90 days – n. (%) | 175/580 (30) | 123/436 (28) | 393/1544 (26) | 0.08 |
| Pneumonia – n. (%) | 40 (6.1) | 55 (12) | 181 (11) | <0.00 |
| NIHSS post intervention –<br>n. (SD) * | 11 (9.6) | 12 (9.7) | 11 (9.5) | 0.18 |
| Procedure time – mean<br>minutes (SD) ^ | 54 (33) | 56 (32) | 57 (33) | 0.14 |
| First-attempt successful<br>(2C-3) – n. (%) | 182/587 (31) | 86/399 (22) | 409/1463 (28) | 0.01 |
| Thrombus in new territory<br>– n. (%) | 29/592 (4.9) | 32/452 (7.1) | 81/1558 (5.2) | 0.24 |
| Distal thrombus – n. (%) | 86/624 (14) | 51/459 (11) | 183/1569 (12) | 0.31 |

\* n=2639, missing in 169 patients; ^ n=2723, missing in 85 patients.
BGC, Balloon guide catheter; Fr, French; mRS, modified Rankin Scale; eTICI, extended Thrombolysis In Cerebral Infarction; ICH, intracranial hemorrhage; NIHSS, National Institutes of Health Stroke Scale.

**Table 3.** Associations between primary and secondary outcomes and the use of a BGC.

| 8-9Fr BGC as first modality | EE | Non-BGC 5-7Fr | Non-BGC 8-9Fr |
| --- | --- | --- | --- |
| mRS at 90 days<sup>*#</sup> | cOR | 0.89 (0.75-1.05) | 0.89 (0.74-1.07) |
|  | acOR | 0.94 (0.78-1.12) | 0.96 (0.79-1.18) |
| mRS 0-1 at 90 days<sup>#</sup> | OR | 0.84 (0.67-1.05) | 0.94 (0.74-1.20) |
|  | aOR | 0.91 (0.71-1.17) | 1.00 (0.76-1.32) |
| mRS 0-2 at 90 days<sup>#</sup> | OR | 1.04 (0.86-1.25) | 0.96 (0.77-1.19) |
|  | aOR | 1.20 (0.95-1.51) | 1.08 (0.83-1.40) |
| Successful reperfusion (eTICI $\geq 2B$)<sup>^</sup> | OR | 0.99 (0.80-1.24) | 0.95 (0.74-1.21) |
|  | aOR | 1.00 (0.80-1.26) | 0.97 (0.75-1.25) |
| Excellent reperfusion (eTICI $\geq 2C$)<sup>^</sup> | OR | 1.10 (0.92-1.33) | 0.98 (0.80-1.21) |
|  | aOR | 1.16 (0.96-1.41) | 1.00 (0.80-1.24) |
| Complete reperfusion (eTICI = 3)<sup>^</sup> | OR | 1.21 (1.00-1.45) | 0.94 (0.76-1.16) |
|  | aOR | **1.26 (1.04-1.53)** | 0.93 (0.75-1.67) |
| Symptomatic ICH | OR | 0.89 (0.60-1.31) | 0.89 (0.57-1.38) |
|  | aOR | 0.91 (0.60-1.37) | 0.81 (0.50-1.31) |
| Ischemic stroke progression | OR | 1.01 (0.72-1.40) | 1.06 (0.74-1.53) |
|  | aOR | 0.98 (0.69-1.39) | 0.97 (0.65-1.44) |
| Pneumonia | OR | **0.53 (0.37-0.76)** | 1.07 (0.78-1.48) |
|  | aOR | **0.49 (0.34-0.72)** | 1.06 (0.75-1.48) |
| Mortality at 90 days | OR | 1.23 (0.99-1.51) | 1.12 (0.88-1.41) |
|  | aOR | 1.14 (0.88-1.47) | 1.00 (0.75-1.33) |
| NIHSS post intervention | $\beta$ | 0.32 (-0.57-1.21) | 0.87 (-0.13-1.87) |
| | aβ | -0.36 (-1.14-0.42) | 0.14 (-0.73-1.02) |
| Improvement on the NIHSS with | OR | 0.97 (0.79-1.18) | **0.79 (0.63-0.98)** |
| ≥4 points | aOR | 1.04 (0.84-1.28) | 0.81 (0.64-1.02) |
| Procedure time | β | -2.92 (-5.92-0.07) | -0.27 (-3.63-3.09) |
|  | aβ | **-3.71 (-6.79- -0.63)** | -2.04 (-5.47-1.38) |
| First-attempt successful | OR | 1.17 (0.96-1.44) | **0.75 (0.58-0.97)** |
| (eTICI ≥2C) | aOR | 1.18 (0.95-1.46) | **0.76 (0.59-0.998)** |
| Thrombus in new territory | OR | 1.03 (0.67-1.59) | 1.40 (0.92-2.14) |
|  | aOR | 0.98 (0.62-1.56) | 1.40 (0.90-2.19) |
| Distal thrombus | OR | 1.20 (0.91-1.58) | 0.97 (0.70-1.34) |
|  | aOR | 1.24 (0.93-1.65) | 0.94 (0.67-1.33) |

\*Common odds ratio for improved mRS score; # mRS is missing in 248 patients; ^ post-eTICI score is missing in 77 patients.
BGC, balloon guide catheter; Fr, French; EE, effect estimate; mRS, modified Rankin Scale; eTICI, expanded Thrombolysis In Cerebral Infarction; ICH, intracranial hemorrhage.

### Safety outcome

sICH was seen in 36 patients (5.5%) in the 5-7Fr non-BGC group, 26 patients in the 8-9Fr non-BGC group (5.5%), and 102 (6.1%) patients in the BGC group (p=0.77). The proportion of patients with pneumonia was lower after the use of a 5-7Fr non-BGC (aOR:0.49, 95%CI:0.34-0.72; Table 3). Distal thrombus was most seen in patients treated with 5-7Fr non-BGC (14%) but did not statically differ from 8-9Fr BGC (12%) (aOR:1.24, 95%CI:0.93-1.65). The rates of thrombus in other territory were similar in all groups, 4.9% in 5-7Fr non-BGC, 7.1% in 8-9Fr non-BGC, and 5.2% in 8-9Fr BGC group (Table 3).

### Subgroup analyses

#### Location of occlusion

There was no significant interaction between location of the occlusion and effect of BGC on the mRS score (p=0.05). Supplemental 2 and 3 show exploratory graphs of the effect of the guide catheters on the mRS based on occlusion location.

#### First line technique

The first-line used technique did interact with the effect of BCG on the mRS score (p<0.001). Patients treated with stent retriever thrombectomy as first-line technique (n=1990) in combination with a 8-9Fr non-BGC had lower chances of better mRS scores compared to the 8-9Fr BGC-SR approach (acOR:0.77, 95%CI:0.59-0.996), and lower first-attempt successful reperfusion rates (aOR:0.52, 95%CI:0.36-0.76) (Table 4).

**Table 4.** Associations between primary and secondary outcomes and the use of a BGC when stent retriever thrombectomy is the first choice of treatment.

| 8-9Fr BGC as first modality | EE | Non-BGC 5-7Fr | Non-BGC 8-9Fr |
| --- | --- | --- | --- |
| mRS at 90 days* | cOR | **0.73 (0.58-0.91)** | **0.76 (0.60-0.98)** |
|  | acOR | 0.81 (0.64-1.03) | **0.77 (0.59-0.996)** |
| mRS 0-1 at 90 days | OR | **0.67 (0.49-0.93)** | 0.79 (0.57-1.11) |
|  | aOR | 0.83 (0.58-1.18) | 0.82 (0.56-1.19) |
| mRS 0-2 at 90 days | OR | 0.90 (0.70-1.17) | 0.81 (0.60-1.07) |
|  | aOR | 1.14 (0.83-1.58) | 0.82 (0.58-1.16) |
| Improvement on the NIHSS with | OR | **0.76 (0.59-0.99)** | **0.72 (0.54-0.95)** |
| ≥4 points | aOR | 0.86 (0.65-1.14) | 0.76 (0.56-1.02) |
| First-attempt successful | OR | 0.80 (0.59-1.07) | **0.50 (0.34-0.72)** |
|  | aOR | 0.83 (0.61-1.13) | **0.52 (0.36-0.76)** |

\*Common odds ratio for improved mRS score.
BGC, balloon guide catheter; Fr, French; EE, effect estimate; mRS, modified Rankin Scale; NIHSS, National Institutes of Health Stroke Scale.

When direct aspiration thrombectomy as first-line technique of treatment was used (n=818), no differences were observed between the BGC and the non-BGC groups on clinical outcomes. In this subgroup, the use of a 5-7Fr non-BGC had higher chances of first-attempt successful reperfusion compared to a BGC (aOR:1.75, 95%CI:1.16-2.63) (Table 5).

**Table 5.** Associations between primary and secondary outcomes and the use of a BGC when direct aspiration thrombectomy is the first choice of treatment.

| 8-9Fr BGC as first modality | EE | Non-BGC 5-7Fr | Non-BGC 8-9Fr |
| --- | --- | --- | --- |
| mRS at 90 days* | cOR | 1.03 (0.77-1.39) | 1.01 (0.73-1.41) |
|  | acOR | 1.02 (0.74-1.41) | 1.21 (0.85-1.72) |
| mRS 0-1 at 90 days | OR | 1.00 (0.68-1.48) | 1.10 (0.73-1.67) |
|  | aOR | 0.98 (0.64-1.55) | 1.25 (0.77-2.02) |
| mRS 0-2 at 90 days | OR | 1.18 (0.84-1.66) | 1.16 (0.80-1.67) |
|  | aOR | 1.32 (0.87-1.99) | 1.55 (0.98-2.47) |
| Improvement on the NIHSS with | OR | 1.37 (0.96-1.96) | 0.97 (0.66-1.42) |
| ≥4 points | aOR | 1.34 (0.90-1.98) | 0.94 (0.61-1.44) |
| First-attempt successful | OR | **1.81 (1.23-2.66)** | 1.22 (0.79-1.89) |
|  | aOR | **1.75 (1.16-2.63)** | 1.22 (0.77-1.95) |

\*Common odds ratio for improved mRS score.
BGC, balloon guide catheter; Fr, French; EE, effect estimate; mRS, modified Rankin Scale; NIHSS, National Institutes of Health Stroke Scale.

## Discussion

In this analysis, there was no overall benefit of the use of a BGC over non-BGC during EVT on functional outcome. However, subgroup analysis suggest higher chances of better functional outcome when a BGC is used in patients treated with stent retriever thrombectomy as first-line technique. Conversely, higher complete reperfusion, shorter procedure times, and less pneumonia rates were in favor of the 5-7Fr non-BGC compared to the BGC.

A recent meta-analysis showed technical, safety, and clinical benefits of BGC, including higher first-attempt successful reperfusion rates, lower distal thrombus rates, and higher mRS 0-2 scores.(3) These results were not confirmed by our study. Contrarily, Supplemental 4 showed higher first attempt successful reperfusion rates when using a non-BGC. Besides, no differences were observed in mRS or distal thrombus rates between the different guide catheters. An explanation for this difference might lie in the fact that our study adjusted for important confounders and used data from 2014 till 2019, whereas the meta-analysis did not adjust and used older data (between 2010 and 2019). In a previous MR CLEAN Registry study, similar analyses were performed on data registered between 2014 and June 2016.

These analyses showed higher reperfusion rates and early improvement of neurologic deficits when using a BGC compared to a non-BGC.(10) Since our study did not showed these benefits after expanding the dataset, this suggests that non-BGCs evolved and that thrombectomy techniques changed (for example the use of intermediate catheters) over the past years. This is in line with Supplemental 5, showing no differences in reperfusion rates and early improvements when using data after June 2016. However, when we take the first-line thrombectomy technique into account, we see comparable results between the subgroup analyses and the results after June 2016 (see Supplemental 6 and 7), with lower chances of better clinical outcomes when using a non-BGC in combination with stent retriever thrombectomy compared to BGC with stent retriever thrombectomy.

Multiple studies report high first pass successful reperfusion rates, especially when using a BGC: Zaidat et al. reported 47.9% and Blasco et al. 45.8%.(7, 11) We reported 51% with 5-7Fr non-BGC, 36% with 8-9Fr non-BGC and 46% with 8-9Fr BGC. Differences can partly be explained by the used technique during EVT. Zaidat et al. and Blasco et al. studied BGC combined with stent retriever thrombectomy, while we included patients treated with either direct aspiration or stent retriever thrombectomy.(7, 11).

Di Maria et al. showed that first-line technique is a potential predictor for first-attempt successful reperfusion, however without differentiating in guide catheter use.(12) We see in our subgroup analyses comparable results regarding the first-line technique as potential predictor, showing a clear interaction between BCG and first-line technique: we observed higher first-attempt successful reperfusion rates when stent retriever thrombectomy as first-line technique was used in combination with a BGC compared to a 8-9Fr non-BGC (Table 4). Additionally, lower odds of better mRS scores were seen in patients treated with 8-9Fr non-BGC and nearly significant in patients treated with 5-7Fr non-BGC in combination with stent retriever thrombectomy compared to BGC. Contrarily, lower chances of first-attempt successful reperfusion were seen in patients treated with direct aspiration thrombectomy and a BGC compared to the 5-7Fr non-BGC. These subgroup analyses might suggest a specific role for a BGC with stent retriever thrombectomy.

Another potential explanation for different rates of first-attempt successful reperfusion is the use of different BGCs. One study showed higher first-attempt successful reperfusion rates after using a FlowGate^2^ catheter compared to a Merci balloon guide catheter.(13) The treating physicians in the MR CLEAN Registry were free to choose the materials of their preference and type or brand of the BGC was not registered properly.

It is assumed that the use of a BGC especially benefits during thrombectomies of proximal occlusions compared to distal occlusions, since the aspiration force is higher around the tip of the BGC compared to distal from the BGC.(14) Although, we may see a trend towards better mRS scores when using a 8-9Fr BGC compared to a 8-9Fr non-BGC for ICA occlusions (Supplemental 3), we found no significant interaction between the location of the occlusion and the use of a BGC on clinical outcome (p=0.05). This may be explained by the fact that the majority of our patients had MCA occlusions.

Some limitations need to be mentioned. First, this was an observational, non-randomized study, with risk of biases, especially since treating physicians were free to choose the guide catheter. The fact that we studied large sample sizes and many treating physicians from all stroke centers in the Netherlands will minimize this effect, although a certain selection bias cannot be ruled out. Second, no distinction could be made between BGCs with and without an inflated balloon. Treating physicians can decide to not inflate the balloon during the procedure, for example when the BGC caused flow arrest already without inflation. In such situation the BGC has the same working mechanism as a non-BGC. Third, although we analyzed the differences between stent retriever and direct aspiration thrombectomy, the combined use of a distal access catheter when using a stent retriever was not registered and is a definite source of heterogeneity in the stentretriever group. Fourth, no proper differentiation was made between short and long sheaths, this data was not reliably registered in the MR CLEAN Registry.

## Conclusion

This large prospective multicenter registry showed no clinical differences between patients treated using a 5-7Fr non-BGC, 8-9Fr non-BGC, or 8-9Fr BGC. However, subgroup analyses suggest that a BCG could be beneficial when a stent retriever is used as first-line technique.

## Data Availability

Due to legislative issues on patient privacy source data will not be made available. On reasonable request to the corresponding author, detailed statistical analyses will be made available.

## Acknowledgments

We thank all the investigators of the MR CLEAN (Multicenter Randomized Controlled Trial of Endovascular Treatment for Acute Ischemic Stroke in the Netherlands) Registry for their effort and contributions.

MR CLEAN Registry investigators:

## Executive committee

Diederik W.J. Dippel^1^; Aad van der Lugt^2^; Charles B.L.M. Majoie^3^; Yvo B.W.E.M. Roos^4^; Robert J. van Oostenbrugge^5,44^; Wim H. van Zwam^6,44^; Jelis Boiten^14^; Jan Albert Vos^8^

## Study coordinators

Ivo G.H. Jansen^3^; Maxim J.H.L. Mulder^1,2^; Robert-Jan B. Goldhoorn^5,6,44^; Kars C.J. Compagne^2^; Manon Kappelhof^3^; Josje Brouwer^4^; Sanne J. den Hartog^1,2,40^; Wouter H. Hinsenveld ^5,6^; Lotte van den Heuvel^1,40^.

## Local principal investigators

Diederik W.J. Dippel^1^; Bob Roozenbeek^1^; Aad van der Lugt^2^; Pieter Jan van Doormaal^2^, Charles B.L.M. Majoie^3^; Yvo B.W.E.M. Roos^4^; Bart J. Emmer^3^; Jonathan M. Coutinho^4^; Wouter J. Schonewille^7^; Jan Albert Vos^8^; Marieke J.H. Wermer^9^; Marianne A.A. van Walderveen^10^; Adriaan C.G.M. van Es^10^; Julie Staals^5,44^; Robert J. van Oostenbrugge^5,44^; Wim H. van Zwam^6,44^; Jeannette Hofmeijer^11^; Jasper M. Martens^12^; Geert J. Lycklama à Nijeholt^13^; Jelis Boiten^14^; Sebastiaan F. de Bruijn^15^; Lukas C. van Dijk^16^; H. Bart van der Worp^17^; Rob H. Lo^18^; Ewoud J. van Dijk^19^; Hieronymus D. Boogaarts^20^; J. de Vries^22^; Paul L.M. de Kort^21^; Julia van Tuijl^21^; Issam Boukrab^26^; Jo P. Peluso^26^; Puck Fransen^22^; Jan S.P. van den Berg^22^; Heleen M. den Hertog^22^; Boudewijn A.A.M. van Hasselt^23^; Leo A.M. Aerden^24^; René J. Dallinga^25^; Maarten Uyttenboogaart^28^; Omid Eschgi^29^; Reinoud P.H. Bokkers^29^; Tobien H.C.M.L. Schreuder^30^; Roel J.J. Heijboer^31^; Koos Keizer^32^; Rob A.R. Gons^32^; Lonneke S.F. Yo^33^; Emiel J.C. Sturm^35,47^, Tomas Bulut^35^; Paul J.A.M. Brouwers^34^; Anouk D. Rozeman^42^; Otto Elgersma^41^, Michel J.M. Remmers^43^; Thijs E.A.M. de Jong^46^.

## Imaging assessment committee

Charles B.L.M. Majoie^3^(chair); Wim H. van Zwam^6,44^; Aad van der Lugt^2^; Geert J. Lycklama à Nijeholt^13^; Marianne A.A. van Walderveen^10^; Marieke E.S. Sprengers^3^; Sjoerd F.M. Jenniskens^27^; René van den Berg^3^; Albert J. Yoo^38^; Ludo F.M. Beenen^3^; Alida A. Postma^6.45^; Stefan D. Roosendaal^3^; Bas F.W. van der Kallen^13^; Ido R. van den Wijngaard^13^; Adriaan C.G.M. van Es^10^; Bart J. Emmer^,3^; Jasper M. Martens^12^; Lonneke S.F. Yo^33^; Jan Albert Vos^8^; Joost Bot^36^; Pieter-Jan van Doormaal^2^; Anton Meijer^27^; Elyas Ghariq^13^; Reinoud P.H. Bokkers^29^; Marc P. van Proosdij^37^; G. Menno Krietemeijer^33^; Jo P. Peluso^26^; Hieronymus D. Boogaarts^20^; Rob Lo^18^;Wouter Dinkelaar^41^; Auke P.A. Appelman^29^; Bas Hammer^16^; Sjoert Pegge^27^; Anouk van der Hoorn^29^; Saman Vinke^20^; Sandra Cornelissen^2^; Christiaan van der Leij^6^; Rutger Brans^6^; Jeanette Bakker^41^; Maarten Uyttenboogaart^28^; Miou Koopman^3^; Lucas Smagge^2^; Olvert A. Berkhemer^1,3,6^; Jeroen Markenstein^3^; Eef Hendriks^3^; Patrick Brouwer^10^; Dick Gerrits^35^.

## Writing committee

Diederik W.J. Dippel^1^(chair); Aad van der Lugt^2^; Charles B.L.M. Majoie^3^; Yvo B.W.E.M. Roos^4^; Robert J. van Oostenbrugge^5,44^; Wim H. van Zwam^6,44^; Geert J. Lycklama à Nijeholt^13^; Jelis Boiten^14^; Jan Albert Vos^8^; Wouter J. Schonewille^7^; Jeannette Hofmeijer^11^; Jasper M. Martens^12^; H. Bart van der Worp^17^; Rob H. Lo^18^

## Adverse event committee

Robert J. van Oostenbrugge^5,44^(chair); Jeannette Hofmeijer^11^; H. Zwenneke Flach^23^

## Trial methodologist

Hester F. Lingsma^40^

## Research nurses / local trial coordinators

Naziha el Ghannouti^1^; Martin Sterrenberg^1^; Wilma Pellikaan^7^; Rita Sprengers^4^; Marjan Elfrink^11^; Michelle Simons^11^; Marjolein Vossers^12^; Joke de Meris^14^; Tamara Vermeulen^14^; Annet Geerlings^19^; Gina van Vemde^22^; Tiny Simons^30^; Gert Messchendorp^28^; Nynke Nicolaij^28^; Hester Bongenaar^32^; Karin Bodde^24^; Sandra Kleijn^34^; Jasmijn Lodico^34^; Hanneke Droste^34^; Maureen Wollaert^5^; Sabrina Verheesen^5^; D. Jeurrissen^5^; Erna Bos^9^; Yvonne Drabbe^15^; Michelle Sandiman^15^; Nicoline Aaldering^11^; Berber Zweedijk^17^; Jocova Vervoort^21^; Eva Ponjee^22^; Sharon Romviel^19^; Karin Kanselaar^19^; Denn Barning^10^; Laurine van der Steen^3^.

## Clinical/imaging data aquisition

Esmee Venema^40^; Vicky Chalos^1,40^; Ralph R. Geuskens^3^; Tim van Straaten^19^; Saliha Ergezen^1^; Roger R.M. Harmsma^1^; Daan Muijres^1^; Anouk de Jong^1^; Olvert A. Berkhemer^1,3,6^; Anna M.M. Boers^3,39^; J. Huguet^3^; P.F.C. Groot^3^; Marieke A. Mens^3^; Katinka R. van Kranendonk^3^; Kilian M. Treurniet^3^; Manon L. Tolhuisen^3,39^; Heitor Alves^3^; Annick J. Weterings^3^; Eleonora L.F. Kirkels^3^; Eva J.H.F. Voogd^11^; Lieve M. Schupp^3^; Sabine L. Collette^28,29^; Adrien E.D. Groot^4^; Natalie E. LeCouffe^4^; Praneeta R. Konduri^39^; Haryadi Prasetya^39^; Nerea Arrarte-Terreros^39^; Lucas A. Ramos^39^; Nikki Boodt^1,2,40^; Anne F.A.V Pirson^5^; Agnetha A.E. Bruggeman^3^; Nadinda A.M. van der Ende ^1,2^, Rabia Deniz^3^, Susanne G.H. Olthuis^5,44^, Floor Pinckaers^6,44^

## List of affiliations

Department of Neurology^1^, Radiology^2^, Public Health^40^, Erasmus MC University Medical Center;

Department of Radiology and Nuclear Medicine^3^, Neurology^4^, Biomedical Engineering & Physics^39^, Amsterdam UMC, location University of Amsterdam;

Department of Neurology^5^, Radiology & Nuclear Medicine^6^, Maastricht University Medical Center+; School for Cardiovascular Diseases Maastricht (CARIM)^44^ and MHeNs School for Mental Health and Neuroscience, Maastricht, the Netherlands^45^;

Department of Neurology^7^, Radiology^8^, Sint Antonius Hospital, Nieuwegein;

Department of Neurology^9^, Radiology^10^, Leiden University Medical Center;

Department of Neurology^11^, Radiology^12^, Rijnstate Hospital, Arnhem;

Department of Radiology^13^, Neurology^14^, Haaglanden MC, the Hague;

Department of Neurology^15^, Radiology^16^, HAGA Hospital, the Hague;

Department of Neurology^17^, Radiology^18^, University Medical Center Utrecht;

Department of Neurology^19^, Neurosurgery^20^, Radiology^27^, Radboud University Medical Center, Nijmegen;

Department of Neurology^21^, Radiology^26^, Elisabeth-TweeSteden ziekenhuis, Tilburg;

Department of Neurology^22^, Radiology^23^, Isala Klinieken, Zwolle;

Department of Neurology^24^, Radiology^25^, Reinier de Graaf Gasthuis, Delft;

Department of Neurology^28^, Radiology^29^, University Medical Center Groningen;

Department of Neurology^30^, Radiology^31^, Zuyderland Medical Center, Heerlen;

Department of Neurology^32^, Radiology^33^, Catharina Hospital, Eindhoven;

Department of Neurology^34^, Radiology^35^, Medisch Spectrum Twente, Enschede, (currently Deventer Hospital^47^);

Department of Radiology^36^, Amsterdam UMC, Vrije Universiteit van Amsterdam, Amsterdam;

Department of Radiology^37^, Noordwest Ziekenhuisgroep, Alkmaar;

Department of Radiology^38^, Texas Stroke Institute, Texas, United States of America;

Department of Neurology^42^, Radiology^41^, Albert Schweitzer Hospital, Dordrecht; Department of Neurology^43^, Radiology^46^, Amphia Hospital, Breda.

